# Investigation of gene-environment interactions in relation to tic severity

**DOI:** 10.1101/2021.05.12.21257005

**Authors:** Mohamed Abdulkadir, Dongmei Yu, Lisa Osiecki, Robert A. King, Thomas V. Fernandez, Lawrence W. Brown, Keun-Ah Cheon, Barbara J. Coffey, Blanca Garcia-Delgar, Donald L. Gilbert, Dorothy E. Grice, Julie Hagstrøm, Tammy Hedderly, Isobel Heyman, Hyun Ju Hong, Chaim Huyser, Laura Ibanez-Gomez, Young Key Kim, Young-Shin Kim, Yun-Joo Koh, Sodahm Kook, Samuel Kuperman, Bennett Leventhal, Marcos Madruga-Garrido, Athanasios Maras, Pablo Mir, Astrid Morer, Alexander Münchau, Kerstin J. Plessen, Veit Roessner, Eun-Young Shin, Dong-Ho Song, Jungeun Song, Frank Visscher, Samuel H. Zinner, Carol A. Mathews, Jeremiah M. Scharf, Jay A. Tischfield, Gary A. Heiman, Andrea Dietrich, Pieter J. Hoekstra

## Abstract

Tourette syndrome (TS) is a neuropsychiatric disorder with involvement of genetic and environmental factors. We investigated genetic loci previously implicated in Tourette syndrome and associated disorders in interaction with pre- and perinatal adversity in relation to tic severity using a case-only (N=518) design. We assessed 98 single nucleotide polymorphisms (SNPs) selected from (I) top SNPs from genome-wide association studies (GWASs) of TS; (II) top SNPs from GWASs of obsessive-compulsive disorder (OCD), attention-deficit/hyperactivity disorder (ADHD), and autism spectrum disorder (ASD); (III) SNPs previously implicated in candidate gene-studies of TS; (IV) SNPs previously implicated in OCD or ASD; and (V) tagging SNPs in neurotransmitter-related candidate genes. Linear regression models were used to examine the main effects of the SNPs on tic severity, and the interaction effect of these SNPs with a cumulative pre- and perinatal adversity score. Replication was sought for SNPs that met the threshold of significance (after correcting for multiple testing) in a replication sample (N = 678). One SNP (rs7123010), previously implicated in a TS meta-analysis, was significantly related to higher tic severity. We found a gene-environment interaction for rs6539267, another top TS GWAS SNP. These findings were not independently replicated. Our study highlights the future potential of TS GWAS top hits in gene-environment studies.

## Introduction

Tourette syndrome (TS) is a childhood onset neuropsychiatric disorder influenced by both genetic and environmental factors. There is clear evidence that implicates both common and rare variants in TS (1,2); however, specific genetic variants only account for a small proportion of total TS disease risk. We investigated the involvement of common SNPs in candidate genes previously implicated in TS and top SNPs from GWAS of TS and comorbid disorders, and found no convincing support for these common variants (3). However, we cannot rule out that these common SNPs might yet confer risk for TS through interaction with environmental factors. Currently, gene-environment (GxE) studies are lacking and only a few small-sampled studies have investigated the genetic etiology of tic severity, suggesting involvement of the dopamine transporter gene (4) and the dopamine receptor D2 gene (5). Unfortunately, no GxE studies have attempted to replicate these initial findings (2). Environmental risk factors such as pre- and perinatal risk factors are also implicated in TS (6); two studies suggested a role for a cumulative score of adverse pre- and perinatal events in TS (7,8).

The aim of the present study was to investigate whether previously implicated SNPs from genome-wide association studies and candidate-gene studies, alone and in interaction with a cumulative pre- and perinatal adversity score, are associated with lifetime tic severity using TS cases recruited by the Tourette International Collaborative Genetics (TIC Genetics) study (9).

## METHODS

### Study subjects

This study included 586 cases (66.7% male; mean age=23.6 years, SD=16.7, range=3-79 years) affected with a chronic tic disorder (458 with TS and 128 with chronic motor or vocal tic disorder) from the ongoing TIC Genetics study (9). As a replication sample, subjects were utilized from the first published TS GWAS (10), including 678 cases (77% male; mean age=18.8 years, SD=14, range=4-78 years) diagnosed with TS (10).

All adult participants and parents of children provided written informed consent along with written or oral assent of their participating child. The Institutional Review Board of each participating site had approved the study.

### Diagnostic assessment

Lifetime worst-ever tic severity (mean = 15.6; SD = 8.22, range 0-30) was assessed based on a modified version of the Yale Global Tic Severity Scale (9). The replication sample included additional items (i.e., number of tics, complexity of tics, and impairment). The mean of both parents’ education level was used as a proxy for socioeconomic status (SES).

### Cumulative pre- and perinatal adversity score

A cumulative pre- and perinatal adversity score (mean = 3.52; SD = 3.42, observed range 0 - 21; previously described in (7)) was constructed from addition of 38 possible adverse events as measured by the self-report or parent-on-child report version of the Modified Schedule for Risk and Protective Factors Early in Development questionnaire (11). Missing values were categorized as absent (coded as 0). The replication sample (10) used the used the same questionnaire (11) in constructing the cumulative perinatal adversity score.

### Selection of single nucleotide polymorphisms

Genetic variants were selected based on a literature review and described in detail elsewhere (3). Briefly, a total of 196 SNPs were assessed: 12 top SNPs from the prior TS GWAS (10,12); 17 top SNPs from GWAS of obsessive–compulsive disorder (OCD; (13)), attention-deficit/hyperactivity disorder (ADHD; (14,15)), and autism spectrum disorder (ASD; (16,17)); 17 SNPs from candidate genes previously implicated (P< 0.05; (3)) in TS; 2 individual candidate SNPs implicated in OCD and one in ASD (3); and 148 tagging SNPs covering seven neurotransmitter-related candidate genes that were either associated with TS, OCD, or ASD (3).

### Genotyping and quality control

Genotyping of 192 SNPs (Table S1) was performed on the Illumina GoldenGate Genotyping Assay for a subset of the cases (N= 464). The remaining cases (N= 122) were genotyped on the HumanOmniExpressExome v1.2 BeadChip genotyping array for a subset of the SNPs (N_SNPs_= 75) available on the Goldengate Assay and four SNPs that were not present on the Goldengate Assay. The total number of SNPs genotyped across both platforms was 196. Standard quality control checks were performed with PLINK (described in detail (3)), which resulted in removal of 10 SNPs. We also removed SNPs with a genotype count less than 20 (N = 80 SNPs) and SNPs located on the X chromosome (N = 8 SNPs) reducing the number of SNPs to 98 (Table S2).

### Statistical analyses

We conducted case-only analyses of tic severity using linear regressions in R (corrected for age, sex, and SES) examining; (I) the main effects of the SNPs on tic severity; and (II) the interaction effect of these SNPs with a cumulative pre- and perinatal adversity score. SNPs were coded as 0 = major allele homozygous (the reference category), 1 = heterozygous, and 2 = minor allele homozygous. Potential confounding due to relatedness of several cases was examined using mixed model analyses with familial relatedness as a random effect.

SNPs were selected from five *a priori* defined groups (Table S2) and we therefore applied correction for multiple testing, first, at the group level by dividing P= 0.05 by the number of SNPs contained within each category; referred to as *P*_*group*_ corrected. To correct for the number of groups tested we further divided the obtained *P*_*group*_ corrected by the number of groups (i.e., five) tested; referred to as the *P*_*all*_. These groups were (I) top SNPs from GWAS of TS, *P*_*group*_ corrected = 0.0071, *P*_*all*_*=* 0.0014 (II) top SNPs from GWAS of OCD, ADHD, and ASD, *P*_*group*_ corrected= 0.0063, *P*_*all*_*=* 0.0013; (III) SNPs previously implicated in candidate-gene studies of TS *P*_*group*_ corrected= 0.005, *P*_*all*_*=* 0.001, (IV) SNPs previously implicated in OCD, or ASD, *P*_*group*_ corrected= 0.0167, *P*_*all*_= 0.0033; and (V) tagging SNPs in neurotransmitter-related candidate genes, *P*_*group*_ corrected =0.0007, *P*_*all*_ =0.0001. For SNPs that met the threshold of multiple testing, replication was sought in an independent sample (10).

## Results

### Sample description

Cases missing clinical or demographic information (N= 68) were excluded, leaving 518 cases eligible for analyses. Results from the mixed model analyses in which a random intercept was included for familial relatedness gave similar results to the models without the random effect (Table S3-4).

### Main effect SNPs

We found a significant association between rs7123010, a top SNP from a GWAS of TS, and tic severity, also after correction for multiple testing (F= 7.99, *P*= 0.0004; Table 1 and Table S1); the AA genotype was positively associated with tic severity.

**Table 1:** Significant results from the main-effect analyses of previously implicated SNPs in relation to lifetime tic severity

| SNP | Position | Chromosome | Gene | Category | F | *P*<sup>a</sup> (SNP) | Main effect in initial sample |  |  |  |  | *P*<sup>a, b</sup><br>(Genotype) | Replication main effect |  |
| --- | --- | --- | --- | --- | --- | --- | --- | --- | --- | --- | --- | --- | --- | --- |
|  |  |  |  |  |  |  | Genotype | N | β | Standard error | T |  | F | *P*<sup>a</sup> |
| rs7123010 | 86341186 | 11 | ME3 | GWAS TS | 7.99 | 0.0004* | GG | 129 |  |  |  |  | 1.98 | 0.14 |
|  |  |  |  |  |  |  | AG | 131 | -1.76 | 0.87 | -2.02 | 0.045 |  |  |
|  |  |  |  |  |  |  | AA | 26 | 3.92 | 1.49 | 2.62 | 0.009 |  |  |

SNP, single nucleotide polymorphism; GWAS, genome-wide association study; TS, Tourette syndrome.
<sup>a</sup>Analyses were corrected for age, sex, and socioeconomic status.
<sup>b</sup>Major allele homozygous genotype was used as the reference genotype.
\*Significant after correcting for multiple testing ( $P_{adj} = 0.0014$ )

### Gene-environment interaction

We found a significant interaction of rs6539267, a top SNP from a TS GWAS (F= 6.80, *P*= 0.001) with the cumulative pre- and perinatal adversity score, also after correction for multiple testing (Table 2; Figure 1); the CC genotype along with a higher number of pre- and perinatal adversities was positively associated with tic severity (Table 2). We found no significant interaction for rs7123010 (F= 0.0197, *P*= 0.98).

**Table 2:** Significant result from the interaction analyses of previously implicated SNPs with a cumulative pre- and perinatal adversity score in relation to lifetime tic severity

| SNP | Position | Chromosome | Gene | Category | F | *P*<sup>a</sup><br>(Interaction SNP) | Interaction in initial sample |  |  |  |  | *P*<sup>a, b</sup><br>(Interaction Genotype) | Replication interaction |  |
| --- | --- | --- | --- | --- | --- | --- | --- | --- | --- | --- | --- | --- | --- | --- |
|  |  |  |  |  |  |  | Genotype | N | Beta | Standard error | T |  | F | *P*<sup>a</sup> |
| rs6539267 | 106785554 | 12 | POLR3B | GWAS TS | 6.80 | 0.001* | TT | 250 |  |  |  |  | 0.43 | 0.65 |
|  |  |  |  |  |  |  | CT | 224 | -0.039 | 0.21 | -1.87 | 0.06 |  |  |
|  |  |  |  |  |  |  | CC | 40 | 1.12 | 0.42 | 2.62 | 0.009 |  |  |

SNP, single nucleotide polymorphism; GWAS, genome-wide association study; TS, Tourette syndrome.
<sup>a</sup>Analyses were corrected for age, sex, and socioeconomic status.
<sup>b</sup>Major allele homozygous genotype was used as the reference genotype.
\*Significant after correcting for multiple testing ( $P_{adj} = 0.0014$ )

**Figure 1:**
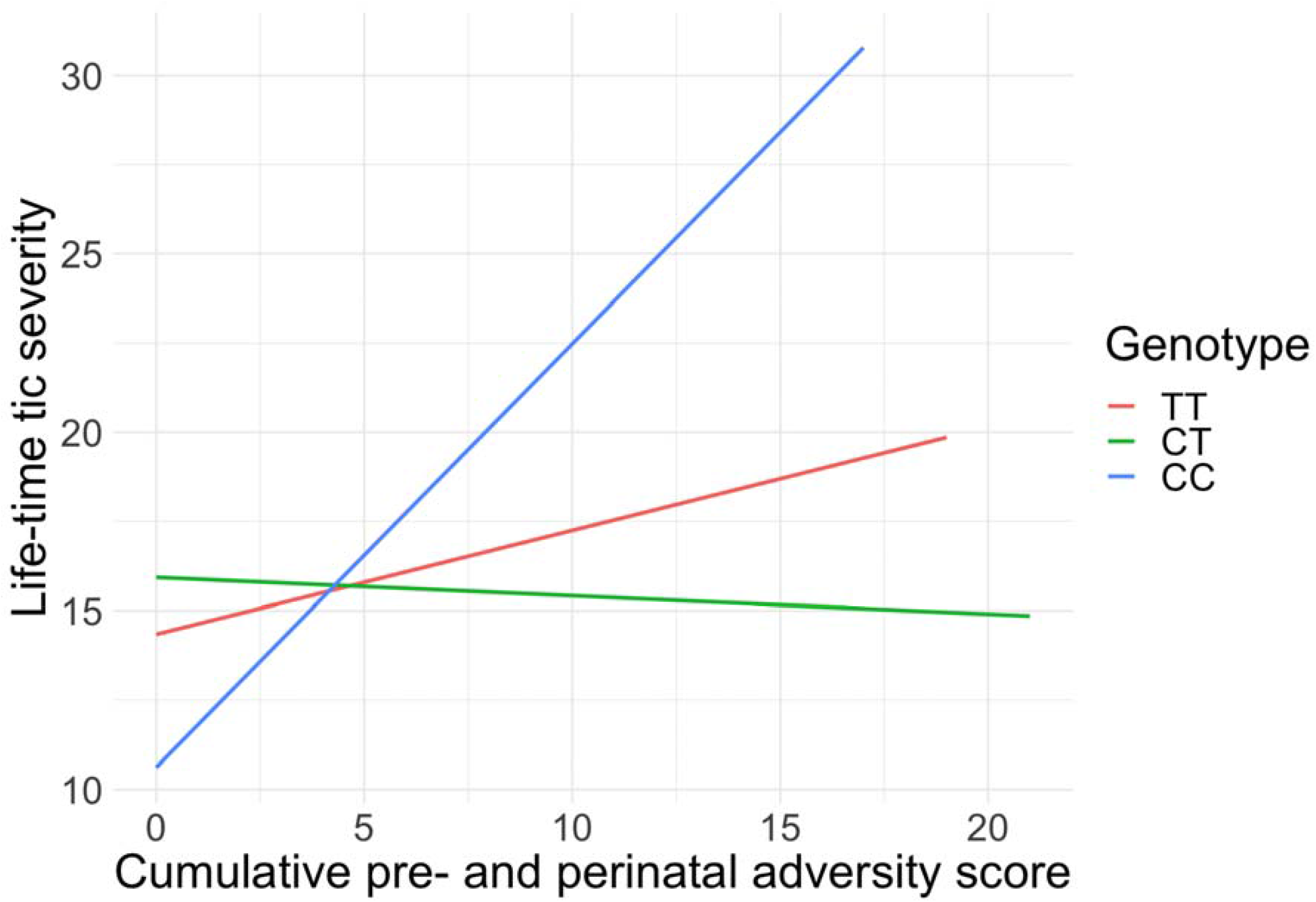
Interaction analyses of rs6539267 with a cumulative pre- and perinatal adversity score in relation to lifetime tic severity.

### Replication rs7123010 and rs6539267

Investigating the main effect of rs7123010 and the interaction between rs6539267 and the cumulative pre- and perinatal adversity score in the replication sample (10) did not show a statistically significant association (F= 1.98, *P*= 0.14) and (F= 1.29, *P*= 0.28), respectively (Table 2).

## Discussion

We investigated whether previously implicated SNPs (i) are associated with lifetime worst-ever tic severity and (ii) might interact with a cumulative pre- and perinatal adversity score previously reported to be associated with TS (7). We report a significant main effect of rs7123010 (a top TS GWAS SNP). We found no evidence for an interaction between rs7123010 and pre- and perinatal adversity. However, we did find a significant interaction between rs6539267 (another top TS GWAS SNP) and pre- and perinatal adversity. We could not confirm these findings in our replication sample (10). This could have been due to the relatively small number of people with the reported effect alleles in the replication sample.

This study benefitted from use of a well-characterized sample, and from the case-only design that has shown to have more power to detect gene-environment interactions than a case-control study (18). Furthermore, using tic severity might have allowed the detection of small effects of SNPs that would have been otherwise missed when investigating caseness; e.g., a significant association for the Dopamine Transporter 1 3’ variable number of tandem repeats has been found in relation to tic severity, but not in relation to the presence of TS (4).

Limitations of this study include the retrospective collection of lifetime tic severity and pre- and perinatal data, although evidence supports accurate maternal long-term recall of the latter (19). Measurement of lifetime tic severity differed across the study and replication samples, yet is not expected to explain current results. Lastly, we cannot exclude that the investigated SNPs might interact with other environmental risk factors, such as life stress or infections.

In conclusion, the findings of this study suggest an association between rs7123010 and tic severity and potential gene-environment interactions of TS GWAS SNP rs6539267 with a cumulative pre- and perinatal adversity score in relation to tic severity. Our study highlights the future potential of common genetic risk variants in gene-environment studies in TS, perhaps through large-scale studies utilizing polygenic scores.

## Data Availability

The clinical data and biomaterials (DNA, transformed cell lines, RNA) are part of a sharing repository located within the National Institute for Mental Health Center for Collaborative Genomics Research on Mental Disorders, USA, and is available to the broad scientific community:
https://www.ncbi.nlm.nih.gov/projects/gap/cgi-bin/study.cgi?study_id=phs001423.v2.p2

## Acknowledgements

We wish to thank the families who have participated in and contributed to this study. We are grateful to New Jersey Center for Tourette Syndrome (NJCTS) for facilitating the inception and organization of the Tourette International Collaborative Genetics (TIC Genetics) study. We would like to thank the members of the TIC Genetics study which are: Yana Bromberg, Lawrence W. Brown, Keun-Ah Cheon, Barbara J. Coffey, Li Deng, Shan Dong, Thomas V. Fernandez, Blanca Garcia-Delgar, Erika Gedvilaite, Donald L. Gilbert, Dorothy E. Grice, Julie Hagstrøm, Tammy Hedderly, Isobel Heyman, Hyun Ju Hong, Chaim Huyser, Laura Ibanez-Gomez, Young Key Kim, Young Shin Kim, Robert A. King, Yun-Joo Koh, Sodahm Kook, Samuel Kuperman, Bennett Leventhal, Marcos Madruga-Garrido, Jeffrey D. Mandell, Athanasios Maras, Pablo Mir, Astrid Morer, Alexander Münchau, Cara Nasello, Kerstin J. Plessen, Petra Richer, Veit Roessner, Stephan Sanders, Eun-Young Shin, Louw Smith, Dong-Ho Song, Jungeun Song, Matthew W. State, Nawei Sun, Frank Visscher, Michael F. Walker, Shuoguo Wang, Jeremy Willsey, Jinchuan Xing, Yeting Zhang, Anbo Zhou, and Samuel H. Zinner. We would also wish to thank the Tourette Association of America International Consortium for Genetics (TAAICG) for providing data that was used as replication of the findings of this study. Members of the TAAICG are: Cathy L. Barr, James R. Batterson, Cheston Berlin, Ruth D. Bruun, Cathy L. Budman, Danielle C. Cath, Sylvain Chouinard, Giovanni Coppola, Nancy J. Cox, Sabrina Darrow, Lea K. Davis, Yves Dion, Nelson B. Freimer, Marco A. Grados, Matthew E. Hirschtritt, Alden Y. Huang, Cornelia Illmann, Robert A. King, Roger Kurlan, James F. Leckman, Gholson J. Lyon, Irene A. Malaty, Carol A. Mathews, William M. MaMahon, Benjamin M. Neale, Michael S. Okun, Lisa Osiecki, David L. Pauls, Danielle Posthuma, Vasily Ramensky, Mary M. Robertson, Guy A. Rouleau, Paul Sandor, Jeremiah M. Scharf, Harvey S. Singer, Jan Smit, Jae-Hoon Sul, and Dongmei Yu.

## Contributors

MA, GAH, PJH, and AD were involved in the organization, design, and execution, critique, and the statistical analysis of the research project. CAM, JMS, DY, and LO were involved in the replication effort of the findings in this study. MA wrote the first draft of the manuscript, which was critically reviewed by JAT, GAH, PJH, and AD who were also involved in the conception of the research project. All authors were involved in the review and critique of the manuscript. All authors have approved the final article.

## Notes

**Conflict of interest** Drs. Mathews and Scharf are on the scientific advisory board of the Tourette Association of America (TAA) and have received travel and grant support from the TAA. Dr. Mathews is also on the scientific advisory board of the International Obsessive-Compulsive Disorder Foundation and the Family Foundation for OCD Research. Dr. Scharf is on the scientific advisory board of the TLC Foundation for Body-Focused Repetitive Behaviors and has received consulting fees from Nuvelution Pharma and Abide Pharmaceuticals. Dr. Coffey is co-Chair of the TAA Medical Advisory Board and has received honoraria from the TAA-CDC partnership She has also received honoraria from the American Academy of Child and Adolescent Psychiatry, Partners Health Care, Harvard Medical School/Psychiatry Academy; consulting fees fromTeva/Nuvelution, and Skyland Trail, and research support from NIMH and Emalex Pharmaceuticals. The remaining authors reported no biomedical financial interest or potential conflict of interest.

**Funding** This research was funded by National Institute of Mental Health (NIMH) grant R01MH092293 (to GAH and JAT) and NJCTS (New Jersey Center for Tourette Syndrome and Associated Disorders; to GAH and JAT). This work was also supported by grants from the Judah Foundation, the Tourette Association of America, National Institute of Health (NIH) Grants NS40024, NS016648, MH079489, MH073250, the American Recovery and Re-investment Act (ARRA) Grants NS040024-07S1; NS16648-29S1; NS040024-09S1; MH092289; MH092290; MH092291; MH092292; R01MH092293; MH092513; MH092516; MH092520; MH071507; MH079489; MH079487; MH079488; and MH079494. Dr. Mir has received grants from the Instituto de Salud Carlos III (PI10/01674, PI13/01461), the Consejería de Economía, Innovación, Ciencia y Empresa de la Junta de Andalucía (CVI-02526, CTS-7685), the Consejería de Salud y Bienestar Social de la Junta de Andalucía (PI-0741/2010, PI-0437-2012, PI-0471-2013), the Sociedad Andaluza de Neurología, the Fundación Alicia Koplowitz, the Fundación Mutua Madrileña and the Jaques and Gloria Gossweiler Foundation. Dr. Morer has received grants from the Fundacion Alicia Koplowitz and belongs to the research group of the Comissionat per Universitats i Recerca del Departmanent d’Innovacio (DIUE) 2009SGR1119. Dr. Münchau has received grants from the Deutsche Forschungsgemeinschaft (DFG: MU 1692/3-1, MU 1692/4-1 and FOR 2698). This study was also supported by a grant from the National Institute for Environmental Health Science (R01 ES021462).

### Competing Interest Statement

Drs. Mathews and Scharf are on the scientific advisory board of the Tourette Association of America (TAA) and have received travel and grant support from the TAA. Dr. Mathews is also on the scientific advisory board of the International Obsessive-Compulsive Disorder Foundation and the Family Foundation for OCD Research. Dr. Scharf is on the scientific advisory board of the TLC Foundation for Body-Focused Repetitive Behaviors and has received consulting fees from Nuvelution Pharma and Abide Pharmaceuticals. Dr. Coffey is co-Chair of the TAA Medical Advisory Board and has received honoraria from the TAA-CDC partnership She has also received honoraria from the American Academy of Child and Adolescent Psychiatry, Partners Health Care, Harvard Medical School/Psychiatry Academy; consulting fees fromTeva/Nuvelution, and Skyland Trail, and research support from NIMH and Emalex Pharmaceuticals. The remaining authors reported no biomedical financial interest or potential conflict of interest.

### Funding Statement

This research was funded by National Institute of Mental Health (NIMH) grant R01MH092293 (to GAH and JAT) and NJCTS (New Jersey Center for Tourette Syndrome and Associated Disorders; to GAH and JAT). This work was also supported by grants from the Judah Foundation, the Tourette Association of America, National Institute of Health (NIH) Grants NS40024, NS016648, MH079489, MH073250, the American Recovery and Re-investment Act (ARRA) Grants NS040024-07S1; NS16648-29S1; NS040024-09S1; MH092289; MH092290; MH092291; MH092292; R01MH092293; MH092513; MH092516; MH092520; MH071507; MH079489; MH079487; MH079488; and MH079494. Dr. Mir has received grants from the Instituto de Salud Carlos III (PI10/01674, PI13/01461), the Consejeria de Economia, Innovacion, Ciencia y Empresa de la Junta de Andalucia (CVI-02526, CTS-7685), the Consejeria de Salud y Bienestar Social de la Junta de Andalucia (PI-0741/2010, PI-0437-2012, PI-0471-2013), the Sociedad Andaluza de Neurologia, the Fundacion Alicia Koplowitz, the Fundacion Mutua Madrilena and the Jaques and Gloria Gossweiler Foundation. Dr. Morer has received grants from the Fundacion Alicia Koplowitz and belongs to the research group of the Comissionat per Universitats i Recerca del Departmanent d'Innovacio (DIUE) 2009SGR1119. Dr. Munchau has received grants from the Deutsche Forschungsgemeinschaft (DFG: MU 1692/3-1, MU 1692/4-1 and FOR 2698). This study was also supported by a grant from the National Institute for Environmental Health Science (R01 ES021462).

### Author Declarations

All adult participants and parents of children provided written informed consent along with written or oral assent of their participating child. The Institutional Review Board of each participating site had approved the study which are: 1. University of Groningen 2. Yale Child Study Center 3. Childrens Hospital of Philadelphia 4. Yonsei University College of Medicine 5. Icahn School of Medicine at Mount Sinai 6. Nathan S. Kline Institute for Psychiatric Research 7. Yulius Mental Health Organization 8. Medizinische Hochschule Hannover Klinik fur Psychiatrie 9. University Hospital Medical Center Hamburg-Eppendorf 10. Hospital Clinic Universitari 11. Cincinnati Childrens Hospital Medical Center 12. Evelina London Childrens Hospital GSTT 13. Great Ormond Street Hospital for Children and UCL Institute of Child Health 14. Hallym University Sacred Heart Hospital 15. De Bascule 16. Amsterdam Academic Medical Center 17. Yonsei Bom Clinic 18. University of California San Francisco 19. Korea Institute for Childrens Social Development 20. Kangbuk Samsung Hospital 21. University of Iowa Carver College of Medicine Iowa City 22. University of Ulm 23. Universidad de Sevilla 24. Erasmus Medical Center-Sophia Childrens Hospital Rotterdam 25. Institut de Investigacions Biomediques August Pi i Sunyer (IDIPABS) and Centro de Investigacion en Red de Salud Mental (CIBERSAM) 26. University of Lubeck 27. University of Copenhagen 28. TU Dresden 29. National Health Insurance Service Ilsan Hospital 30. Altrecht Institute for Mental Health 31. Admiraal De Ruyter Ziekenhuis 32. University of Washington Seattle

